# Targeted encouragement of GP consultations for possible cancer symptoms: randomised controlled trial

**DOI:** 10.1101/2020.06.09.20126540

**Authors:** Jean-Pierre Laake, Daniel Vulkan, Samantha L Quaife, William T Hamilton, Tanimola Martins, Jo Waller, Dharmishta Parmar, Peter Sasieni, Stephen W Duffy

**Author notes:** Corresponding Author: Daniel Vulkan, Queen Mary University of London, Centre for Cancer Prevention, Wolfson Institute of Preventive Medicine, Charterhouse Square, London EC1M 6BQ. Daniel Vulkan and Jean-Pierre Laake contributed equally and are joint first authors of this work. Peter Sasieni and Stephen Duffy contributed equally and are joint last authors of this work.

## Abstract

**Background:** For some common cancers, survival is lower in the UK than in comparable high-income countries.

**Aim:** To assess the effectiveness of a targeted postal intervention (promoting awareness of cancer symptoms and earlier help-seeking) on patient consultation rates.

**Design and Setting:** A two-arm randomised controlled trial (RCT) of adult patients registered at 23 general practices in England.

**Method:** Adult patients who had not consulted their general practice in the previous 12 months and had at least two other risk factors for late presentation with cancer were randomised to intervention and control arms. The intervention consisted of a mailed letter. The primary outcome was number of consultations at the practice in the six months subsequent to mailing of the intervention. All patients with outcome data were included in the intention-to-treat analyses. The trial was registered prospectively on the International Standard Randomised Controlled Trial Number (ISRCTN) registry (ISRCTN95610478).

**Results:** 1,513 patients were individually randomised to the intervention (n=783) and control (n=730) arms, between November 2016 and May 2017. Outcome data was available for 749 and 705 patients respectively. There was a significantly higher rate of consultation in the intervention arm: 436 consultations compared to 335 in the control arm (RR = 1.40, 95% CI 1.11-1.77, p=0.004). However, there was no difference in the numbers of patients consulting.

**Conclusion:** Targeted interventions of this nature can change behaviour. There is a need to develop interventions which can be more effective at engaging patients with primary care.

**HOW THIS FITS IN:** Later stage of cancer diagnosis is associated with poorer survival and may arise from low symptom awareness and from delays in presenting to primary care. Population-wide campaigns to increase awareness and encourage help-seeking have shown mixed results in terms of stage at diagnosis and numbers of primary care consultations. This randomised controlled trial was targeted at a population whose circumstances would suggest that should they develop cancer, they are at increased risk of being diagnosed with later stage disease. This study demonstrates targeted interventions of this nature can change consultation behaviour of those who are likely to benefit most from earlier symptomatic presentation.

## INTRODUCTION

For some common cancers, survival in the UK is lower than in comparable high-income countries.^1^ It is thought that this is due largely to later stage disease at diagnosis. This may be partly related to system delays following presentation, some of which have increased over the last decade in England.^2^ It may also arise from low symptom awareness, negative beliefs about cancer and reluctance to ‘waste the doctor’s time’,^3,4^ all of which could extend the patient interval.^5^ There is a positive association between levels of awareness of cancer and survival.^6^ In addition, there remain geographic and socioeconomic inequalities in cancer survival, although the gap has narrowed somewhat in recent years.^7,8^

Several campaigns, including ‘Be Clear on Cancer,’ have aimed to raise the level of symptom awareness and encourage help-seeking for symptoms.^9^ Some have shown encouraging results with respect to diagnosis of cancer at an earlier stage.^10,11^ Others have shown an increase in consultations and referrals, but no effect on diagnoses of cancer or stage of disease at diagnosis.^12^ This gives rise to the concern that population-level, mass media campaigns may increase consultations among the ‘worried well’, rather than reaching the population most in need of earlier presentation.

This led to speculation that targeted rather than whole-population symptom awareness interventions might be more effective in reducing the patient interval, and at improving stage at symptomatic presentation. Targeting in this context is at those whose circumstances or lifestyle imply that, should they develop cancer, they are at increased risk of diagnosis at a later stage. This population includes those of lower socioeconomic status, smokers, those with chronic co-morbidities, certain ethnic groups, and those who tend to use emergency services rather than primary care.^13-17^ We carried out a randomised controlled trial (RCT) to test the effectiveness of a postal intervention, targeted towards the aforementioned groups, designed to promote awareness of cancer symptoms and earlier help-seeking at general practice.

## METHODS

### Study design and setting

Here we report on WELCOME-GP (Writing to Encourage Late Consultation Outpatients to Make Engagement with their GP), a two-arm RCT examining the effectiveness of a postal intervention designed to promote awareness of six cancer ‘red flag’ symptoms, and to allay fears of wasting the doctor’s time. The intervention was targeted at a population that had not attended their general practice in the last year, with factors that suggest this may be incongruous with their clinical need and with attributes known to be risk factors for late stage presentation of cancer.^13-17^ A total of 24 practices from three areas of England (the North East, Greater Manchester and London) agreed to take part, with one subsequently withdrawing.

### Patients

Patients were eligible for inclusion if they were aged 50-84, registered with a participating practice, had not had a consultation at their registered practice in the last 12 months and satisfied at least <u>two</u> of the following:

- Low socioeconomic status (two lowest 2015 Index of Multiple Deprivation (IMD) quintiles based on post code^18^)
- Missed last breast, bowel, cervical cancer or abdominal aortic aneurism screen
- History of use of emergency or out-of-hours services instead of primary care
- Missed last appointment for chronic disease monitoring / management
- Living alone (indexed by being the only person registered with the practice at their address) as a marker of social isolation
- Smoker (ever)

However, we were unable to identify with confidence patients who had used emergency or out-of-hours services from the practice databases. Therefore, no patients were recruited on this basis.

Persons were ineligible if they already had a diagnosis of cancer, if the GP considered recruitment inappropriate in view of the patient’s state of health or cognitive capacity, or if the GP felt that the patient would not wish to be included in the research.

### Randomisation

Sample size calculations are described in Supplementary Appendix 1. Each practice was provided with a computer-generated randomisation schedule provided by the research team. Practice staff identified eligible patients and randomly allocated patients to the intervention or to usual care; there was no concealment of allocation. A total of 1,513 patients were randomised using simple 1:1 randomisation (Figure 1).

**Figure 1.**
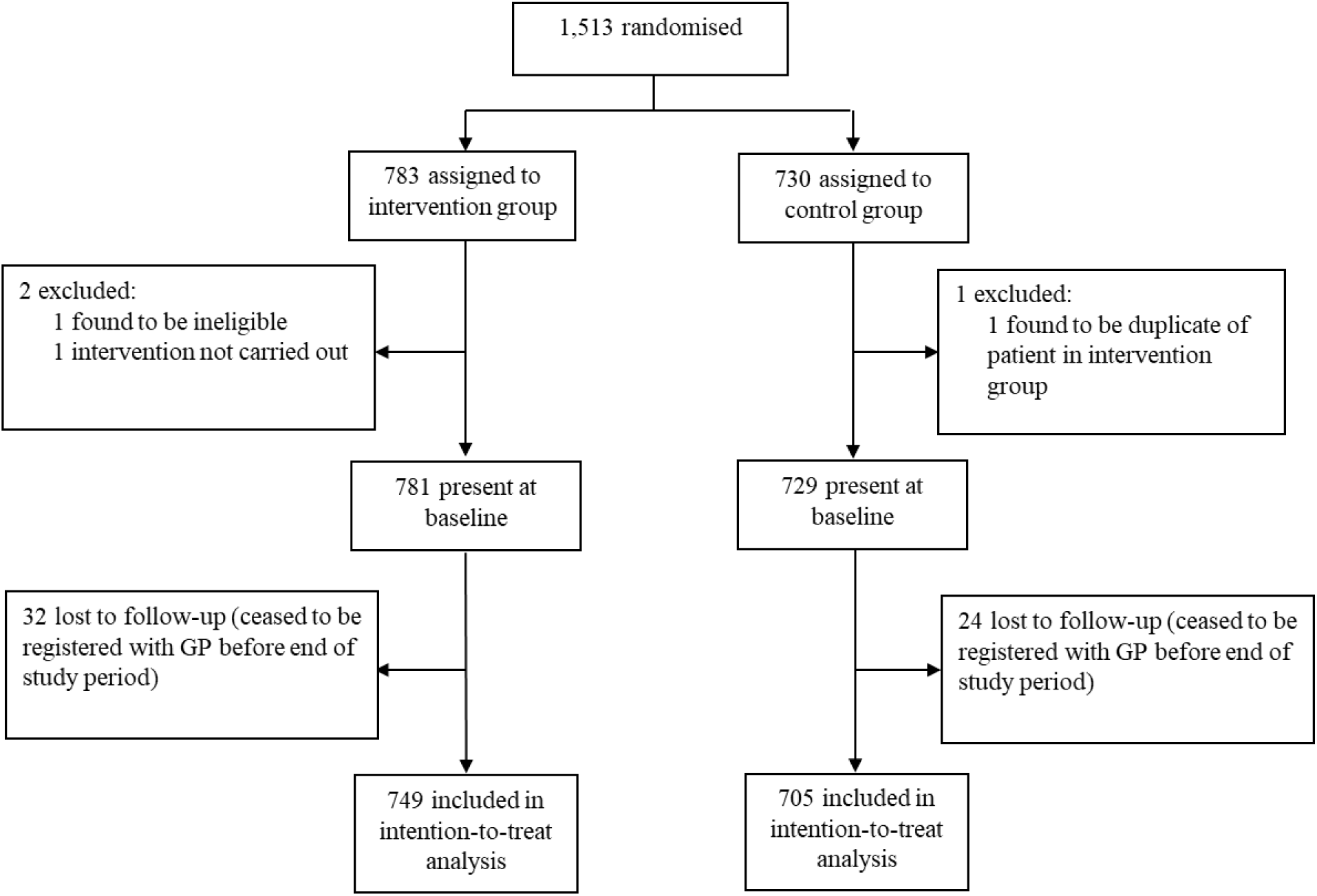
WELCOME-GP Trial profile (adapted from CONSORT template)

At the end of the six month follow-up period, staff at each practice reported the number of consultations by each patient who continued to be registered throughout the period plus: the month of the consultation, the reason for the consultation, whether or not there were any investigations or referrals to secondary care.Intention-to-treat analyses were carried out for all eligible patients for whom data was available at follow-up.

Due to the nature of the intervention, it was necessary that patients in both groups were unaware that they were taking part in a research study. Practice staff and the data analysis team were not blinded to the allocation.

### Intervention

The Model of Pathways to Treatment provides a framework for understanding the pathways between an individual first detecting a bodily change and undergoing treatment^19^ beginning with ‘appraisal’ and ‘help-seeking’ intervals. Crucially, the individual must believe there to be a reason to present with their symptom, often motivated by heuristics such as concern about a worsening symptom or its interference with their daily activities.^20^ The present intervention can be contextualised within these intervals, in that it was intended to prompt appraisal through symptom awareness, provide individuals with a ‘cue to action’ for visiting their GP, and use messaging to counteract known attitudinal barriers to help-seeking. Furthermore, the personalised, GP-led approach drew on the success of primary care endorsement in the cancer screening context.^21,22^

The intervention consisted of a letter, signed by one of the GPs at the patient’s practice, noting that the recipient had not been seen at the practice for some time and reassuring the recipient that if he or she should consult them with any symptoms, this would not be considered wasting the doctor’s time. The letter also drew attention to an enclosed leaflet detailing six symptoms potentially suspicious for cancer: blood in urine; blood in stool; persistent cough; haemoptysis; difficulty swallowing; and unexplained weight loss. These symptoms were chosen as they feature in the National Institute for Health and Care Excellence recommendations for referral for suspected cancer.^23^ The letter and leaflet are given in Supplementary Appendix 2 and Supplementary Appendix 3 respectively. The first practice mailed the intervention materials to all patients allocated to the intervention arm on 11 November 2016, and the last on 31 May 2017.

### Outcomes

The primary endpoint was the rate of consultations at the patient’s registered practice in the six months following randomisation (i.e. the date that letters were mailed). Secondary endpoints were total general practice activity (consultations, referrals, investigations and diagnoses) and the use of emergency and out-of-hours services in the study and control groups.

As noted above, it could not be established with confidence whether patients had made use of emergency services, so this particular analysis was not possible. It also became clear during the course of the study that it would not be possible to collect data on subsequent diagnoses, as these may have occurred some time after the end of the six month follow-up.

Several additional analyses were carried out. The primary and secondary analyses were repeated, considering only those consultations which had taken place for one or more of the six symptoms identified in the leaflet. A further comparison was carried out of the number of consultations which took place during the same calendar month as randomisation.

### Statistical analyses

All randomised patients for whom follow up data was available were included in the intention-to-treat analyses. Data were analysed by zero-inflated Poisson regression,^23^ a technique which takes account of the large number of persons with no consultations at all during the six-month observation period. Relative risks (RR) of consultation with 95% confidence intervals (CI) were calculated. Robust variance estimators were used, allowing for clustering by person. Stata version 15.1 was used for data analysis.

## RESULTS

Table 1 shows the age, sex, eligibility criteria satisfied and region of residence of the patients randomised. Figure 1 shows the trial profile. Of the 1,513 randomised, 1,454 (96%), (749 in the intervention arm and 705 in the control arm) completed six months within the practice in which they were randomised, and therefore had a primary endpoint for analysis.

**Table 1.** Basic description of the study population.

| Table 1. Basic description of the study population |  |  |  |
| --- | --- | --- | --- |
| Factor | Category | Intervention no. (%) | Control no. (%) |
| Patients |  | 781 | 729 |
| Age | <50 | - | - |
|  | 50-59 | 621 (79.5) | 557 (76.4) |
|  | 60-69 | 104 (13.3) | 113 (15.5) |
|  | 70+ | 56 (7.2) | 59 (8.1) |
| Sex | Male | 546 (69.9) | 500 (68.6) |
|  | Female | 235 (30.1) | 229 (31.4) |
| Low socioeconomic status | No | 115 (14.7) | 118 (16.2) |
|  | Yes | 666 (85.3) | 611 (83.8) |
| Missed last screening appointment | No / not applicable | 565 (71.6) | 522 (72.3) |
|  | Yes | 216 (28.4) | 207 (27.7) |
| Missed last chronic disease monitoring appointment | No | 628 (80.4) | 610 (83.7) |
|  | Yes | 153 (19.6) | 119 (16.3) |
| Living alone | No | 394 (50.5) | 381 (52.3) |
|  | Yes | 387 (49.5) | 348 (47.7) |
| Smoker | No | 84 (10.8) | 81 (11.1) |
|  | Yes | 697 (89.2) | 648 (88.9) |
| Region | North East England | 241 (30.9) | 221 (30.3) |
|  | Greater Manchester | 280 (35.9) | 273 (37.4) |
|  | Greater London | 260 (33.3) | 235 (32.2) |

## STATISTICS

### Primary analyses

Table 2 shows general practice consultations and onward referrals in the two trial arms, and the reasons given for consultations. There was a significantly higher rate of consultation in the intervention arm compared to the control arm (436 vs. 335) (RR = 1.40, 95% CI 1.11-1.77, p=0.004), but no difference between arms in the likelihood of individuals consulting (odds ratio =0.92, 95% CI 0.72-1.18, p=0.53).

**Table 2.**
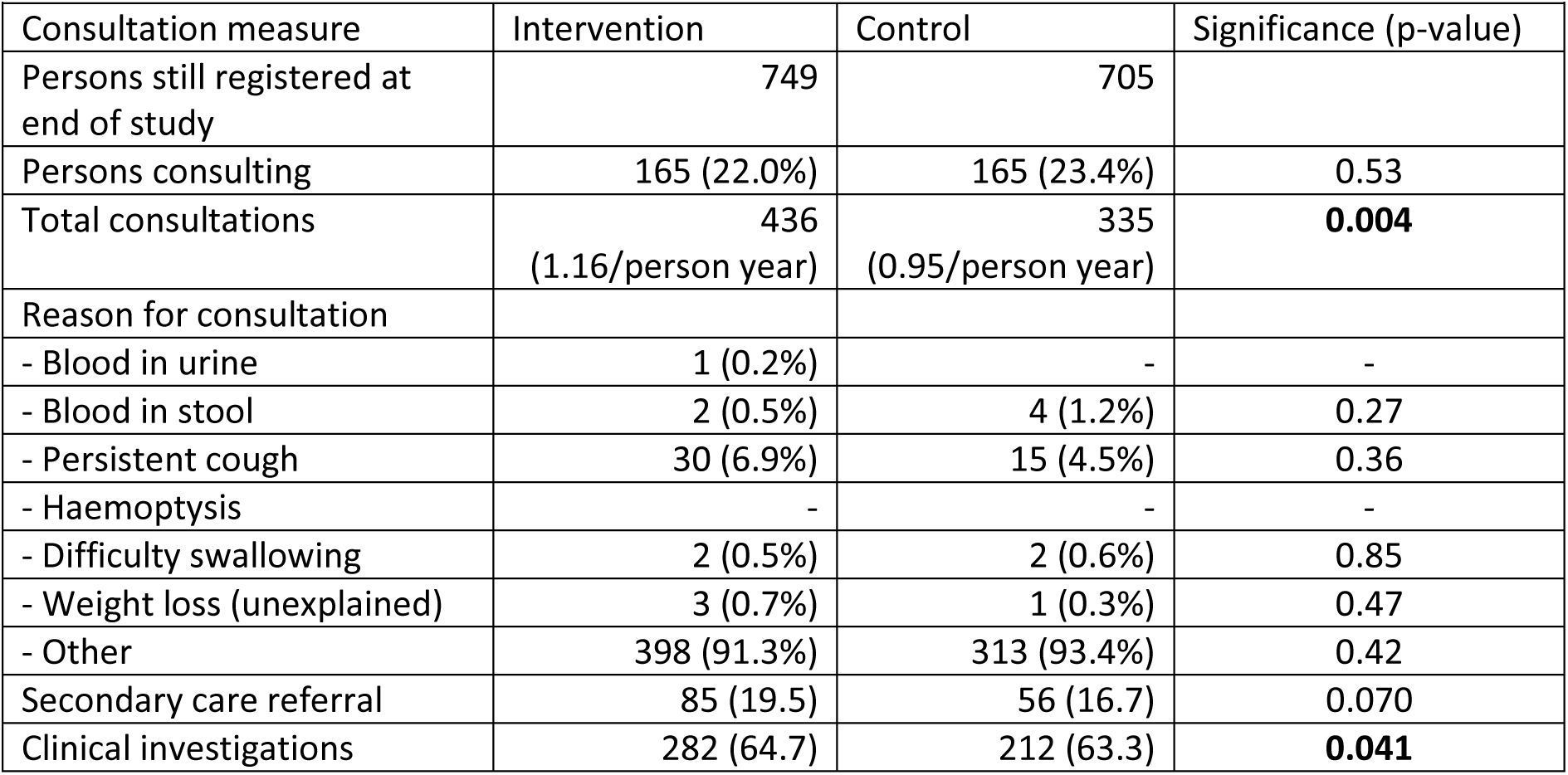
Consultations, reasons for consultation, onward referrals to secondary care, and clinical investigations, in the intervention and control groups

|  |  |  |  |
| --- | --- | --- | --- |
| Consultation measure | Intervention | Control | Significance (p-value) |
| Persons still registered at end of study | 749 | 705 |  |
| Persons consulting | 165 (22.0%) | 165 (23.4%) | 0.53 |
| Total consultations | 436<br>(1.16/person year) | 335<br>(0.95/person year) | **0.004** |
| Reason for consultation |  |  |  |
| - Blood in urine | 1 (0.2%) | - | - |
| - Blood in stool | 2 (0.5%) | 4 (1.2%) | 0.27 |
| - Persistent cough | 30 (6.9%) | 15 (4.5%) | 0.36 |
| - Haemoptysis | - | - | - |
| - Difficulty swallowing | 2 (0.5%) | 2 (0.6%) | 0.85 |
| - Weight loss (unexplained) | 3 (0.7%) | 1 (0.3%) | 0.47 |
| - Other | 398 (91.3%) | 313 (93.4%) | 0.42 |
| Secondary care referral | 85 (19.5) | 56 (16.7) | 0.070 |
| Clinical investigations | 282 (64.7) | 212 (63.3) | **0.041** |

Onward referral rates were higher in the intervention arm, but not significantly 85 (19.5% of consultations) vs. 56 (16.7%) (RR = 1.44, 95% CI 0.97-2.14, p=0.070). There was a significant difference in the number of clinical investigations carried out as a result of consultations, with 282 (64.7%) in the intervention arm compared to 212 (63.3%) in the control arm (RR = 1.34, 95% CI 1.01-1.77, p=0.041).

### Additional analyses

The number of consultations for the symptoms described in the leaflet was higher in the intervention than in the control arm (38 vs. 22), although this was not significant (RR=1.74, 95% CI 0.81-3.74, p=0.16). Onward referral in relation to symptoms in the leaflet did not differ by arm (8 vs. 8; RR=0.88, 95% CI 0.25-3.07, p=0.84). There were more subsequent investigations in the intervention arm for symptoms in the leaflet (34 vs. 19) but this difference was also not significant (RR=1.94, 95% CI 0.90-4.21, p=0.093). There were no significant differences between the intervention and control arms with regard to the reasons given for consultation.

Table 3 shows the number of consultations in the calendar month of randomisation and in subsequent months. Table 4 shows the month in which the first consultation took place. In the intervention arm, there were significantly more consultations during the calendar month of randomisation than in the control arm, at 16 vs. 8 (RR=2.50, 95% CI 1.07-5.86, p=0.035). However, there was no significant difference, between the arms, in the month of first consultation (0.3 months earlier in intervention arm, 95% CI -0.7-0.1, p=0.10). Whilst the date of randomisation in each practice was known, exact dates of consultations were not. Sensitivity analyses determined that the date of the month on which randomisation occurred did not have any impact on the statistical significance of these results.

**Table 3.**
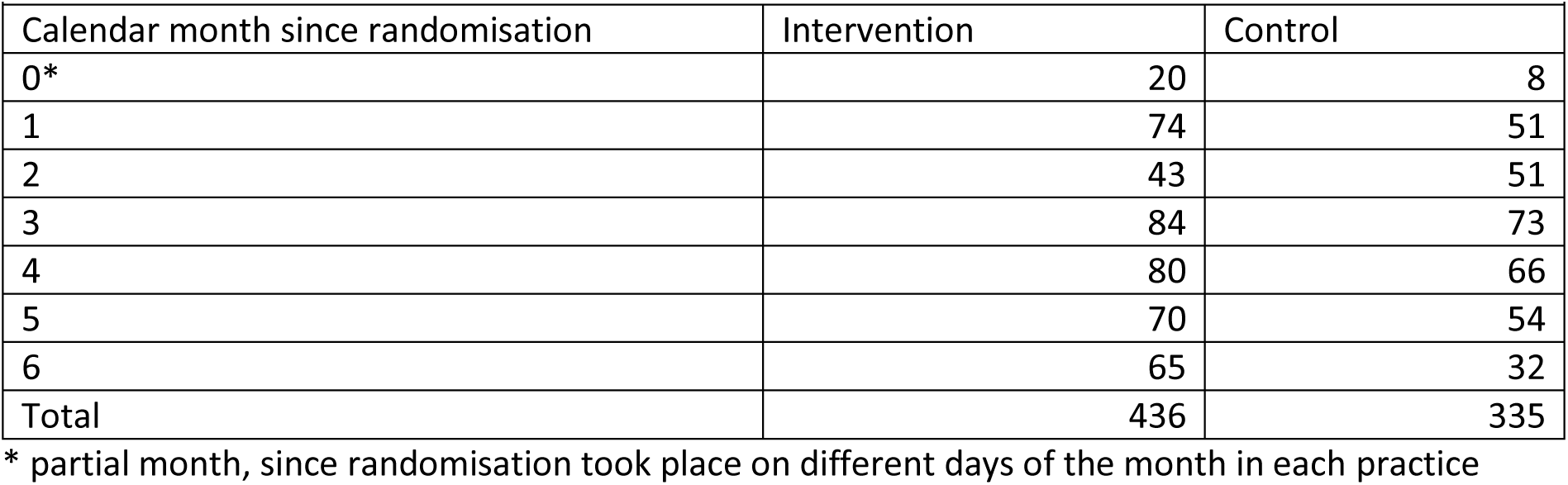
Number of consultations in each calendar month since randomisation, in the intervention and control groups

|  |  |  |
| --- | --- | --- |
| Calendar month since randomisation | Intervention | Control |
| 0* | 20 | 8 |
| 1 | 74 | 51 |
| 2 | 43 | 51 |
| 3 | 84 | 73 |
| 4 | 80 | 66 |
| 5 | 70 | 54 |
| 6 | 65 | 32 |
| Total | 436 | 335 |

\* partial month, since randomisation took place on different days of the month in each practice

**Table 4.** Calendar month since randomisation in which first consultation took place, in the intervention and control groups

|  |  |  |
| --- | --- | --- |
| Calendar month since randomisation | Intervention | Control |
| 0* | 16 | 8 |
| 1 | 37 | 34 |
| 2 | 20 | 25 |
| 3 | 40 | 31 |
| 4 | 23 | 28 |
| 5 | 15 | 24 |
| 6 | 14 | 15 |
| Total | 165 | 165 |
| No consultations within six months | 584 | 540 |

\* partial month, since randomisation took place on different days of the month in each practice

## DISCUSSION

### Summary

This trial took place in a population which was interacting less with primary care and which had factors associated with a lesser tendency to help-seek and late presentation of cancer.^13-17,25,26^ The results demonstrate a targeted primary care-based intervention can change consulting behaviour in this population, but not necessarily in the way expected. In the intervention group there were significantly more general practice consultations than in the control group, but there was no increase in the number of ***persons*** consulting.

There were twice as many consultations for the six ‘red flag’ cancer symptoms detailed in the leaflet in the intervention group than the control group, but this was not statistically significant. However, given the relatively small proportion of the population targeted in this trial, it is possible that were more primary care providers incentivised to deliver such a targeted intervention, this might result in statistically and clinically significant numbers of increased primary care consultations for cancer ‘red flag’ symptoms and referrals. Irrespective of the symptom(s) patients presented with, if the increase in consultation rates observed amongst the target patient group in this trial were to be sustained this may result in earlier diagnoses of cancer. This is because GPs generally enquire about ‘red flag’ symptoms when patients present with other lower-risk possible cancer symptoms. Increased consultation rates also provide GPs with an opportunity to promote primary and secondary prevention of cancer and other disease by supporting patients to attend non-symptomatic screening,^29^ to manage other chronic conditions more effectively, and to adopt healthier behaviours through initiatives such as Making Every Contact Count.^28^ These are activities GPs already routinely perform for patients who present frequently, which may partially explain inequalities in outcomes in groups that access services less.

### Strengths and limitations

There were several strengths to this trial. Firstly we were able to recruit a relatively large sample of patients from typically hard to reach groups registered at both inner-city and rural practices across England, which gives us confidence that the results can be applied to the national population. Secondly, this intervention was relatively cheap and was acceptable to most practices approached. Thirdly, the unusual design of the study, with no study specific prior informed consent process, meant the trial was not limited by selection bias for patients who are most interested in and prepared to alter behaviour. While we were able to measure presenting behaviour, a limitation of this study was that we were unable to assess the duration of effect and the impact on clinical end-points. Another limitation was the ambitious concept of changing behaviour of a particularly difficult to reach group with a single postal communication. Therefore, we do not know whether a second letter would increase effect or if there is a ‘dose response’ to this intervention. There are several stages in the process of cancer awareness, willingness to seek help, referral and diagnosis, and we considered we were not at the stage of sufficient knowledge to assess the impact of a psychosocial intervention on cancer diagnoses.

### Comparison with existing literature

The intervention is only a partial solution to the problem of incongruous consultation behaviour in primary care, particularly bearing in mind pressures on capacity. The targeted promotion of cancer awareness and help-seeking for symptoms may have a contribution to make in increasing and modifying help-seeking behaviours and in reducing inequity in access to care and the inequality in cancer survival, in a way that mass campaigns may not.^9^

### Implications for research and/or practice

Similar interventions may have applications in mitigating other health inequalities by facilitating more equitable access to other areas of primary care (such as pharmacies and psychological wellbeing services).^30,31^ Other applications may include facilitating more equitable access to support for health and wellbeing promotion and for the primary, secondary and tertiary prevention of disease, such as recent letters advising those at increased risk to ‘shield’ from COVID-19 by staying at home and signposting recipients to the Every Mind Matters website.^32,33^

During the COVID-19 pandemic there have been fewer referrals to secondary care, and fewer diagnoses of cancer. Patients are balancing the risks of infection with the risks of delayed diagnosis. This unmet need may in part by met by initiatives as described here, though such an intervention will have to be timed to match re-opening of diagnostic services.

This trial demonstrates the potential of an inexpensive targeted postal symptom awareness intervention for altering consultation behaviour and reducing barriers to help-seeking in general practice. There is a need to evolve similar interventions with the potential to support a wider range of potential patients. This is particularly relevant in the UK given the commitment of the NHS to providing equitable care.^34^

## Data Availability

The relevant anonymised patient level data will be made available on reasonable request to the corresponding author.

## SUPPLEMENTARY APPENDICIES

Supplementary Appendix 1. Sample size calculations

Supplementary Appendix 2. Intervention letter

Supplementary Appendix 3. Intervention leaflet

## ADDITIONAL INFORMATION

### Ethical Approval

Ethical approval was granted by the West of Scotland Research Ethics Service (WoSREC) (reference 16/WS/0110). The study was approved by the Health Research Authority (HRA) (reference 201992). Due to the nature of the study it was necessary that patients allocated to both groups were not aware that they were taking part in the study, as such contacting patients to obtain prior informed consent would have defeated the object of the study. The study was performed in accordance with the Declaration of Helsinki. Ethical approval was received to carry out the study without seeking prior informed consent from patients.

## Acknowledgements

The authors would like to thank staff at the participating practices for carrying out the randomisation, delivery of the intervention, data collection and data extraction. The authors also thank the CRN North West London, CRN Greater Manchester, CRN North East and North Cumbria and CRN North Thames for their assistance in recruiting and accessing practices. We thank Greg Rubin and Rosalind Raine for helpful discussion.

## Authors’ contributions

JPL & DV drafted the manuscript, developed the protocol, managed the trial (HRA and ethical approval, supervised training of practice staff for randomisation, delivery of the intervention, data collection and data extraction) and contributed equally to this paper. SWD and PS conceived the idea for this trial, the analysis plan, supervised the data analysis and created the study protocol. TM designed the intervention materials, which were further developed by WH, JPL, SD, SLQ and JW. All authors revised the manuscript for important intellectual content and approved the final version for submission. DP was responsible for data processing and informatics.

DV & SW affirm that the manuscript is an honest, accurate and transparent account of the study being reported, that no important aspects of the study have been omitted, and that all discrepancies from the study as planned and registered have been explained.

## Trial Registration

This trial was registered prospectively on the International Standard Randomised Controlled Trial Number (ISRCTN) registry (ISRCTN95610478).

## APPENDIX S1 Sample size calculations

The average rate of general practitioner (GP) consultation in the age group 50-84 is around 8 per person per year. It was assumed that, in the target population without intervention, the rate would be only 0.8 per year, or 40 consultations per 100 people over a six-month period. If the intervention were to increase the rate of consultation from 40 per 100 to 55 per 100 (i.e. 15 percentage points), and if the number of consultations were independent Poisson distributed, 547 persons per group would confer 90% power for the comparison (5% significance level, two-sided testing). Because consultations are not necessarily independent, it was proposed to adopt, as a fail-safe, 700 subjects per group, a total of 1,400, with the intention to recruit 70 subjects from each of 20 general practices, allocated 1:1 between the intervention and usual care within each practice.

## APPENDIX S2 Intervention letter

**XXXXXXXXXXX NHS Health Centre**

XXXXXXX Road, Town, County. POST CODE

Patient ID number <Patient ID>

<Patient Given Name> < Patient Family Name >

<Preferred Mailing Address>

<Preferred Mailing Address >

<LOCALITY> <STATE> <POSTCODE>

Dear < Patient Given Name> < Patient Family Name >

I hope this finds you well. We are writing to patients we have not seen for some time to ask them to make an appointment if they have any concerns about their health.

I enclose a leaflet about symptoms which are usually harmless but can sometimes be a sign of something more serious. We recommend patients with any of the symptoms listed in the leaflet come to the surgery to discuss them with their doctor.

I know many people worry about wasting the doctor’s time. However, I would like to assure you that if you do come to see us with any of these symptoms you would NOT be wasting our time.

Please call us on **XXXXXXXXXXX** if you think you might have any of these symptoms and would like to book an appointment to see your doctor. Please also consult us anytime about any aspect of your health which is worrying you. Our patients are important to us.

Thank you for taking the time to read this letter and the enclosed leaflet.

Yours sincerely,

DR NAME SURNAME

XXXXXXXXXXX NHS Health Centre

<DATE>

## APPENDIX S3 Intervention leaflet

**What happens next?**

If you have a symptom, please call your doctor to make an appointment. We’re here to help.

It’s easier for your doctor to make an assessment of your condition if they can see you in person but if it is not convenient to attend you should ask us if we can arrange a telephone appointment.

**Further information**

If you would like to know more about cancer or the symptoms covered in this leaflet, please visit the

CRUK website:

http://www.cancerresearchuk.org/about-cancer/cancer-symptoms

V1.0 29.03.16

**Your doctor will be happy to see you (we know some people worry about wasting the doctor’s time)**

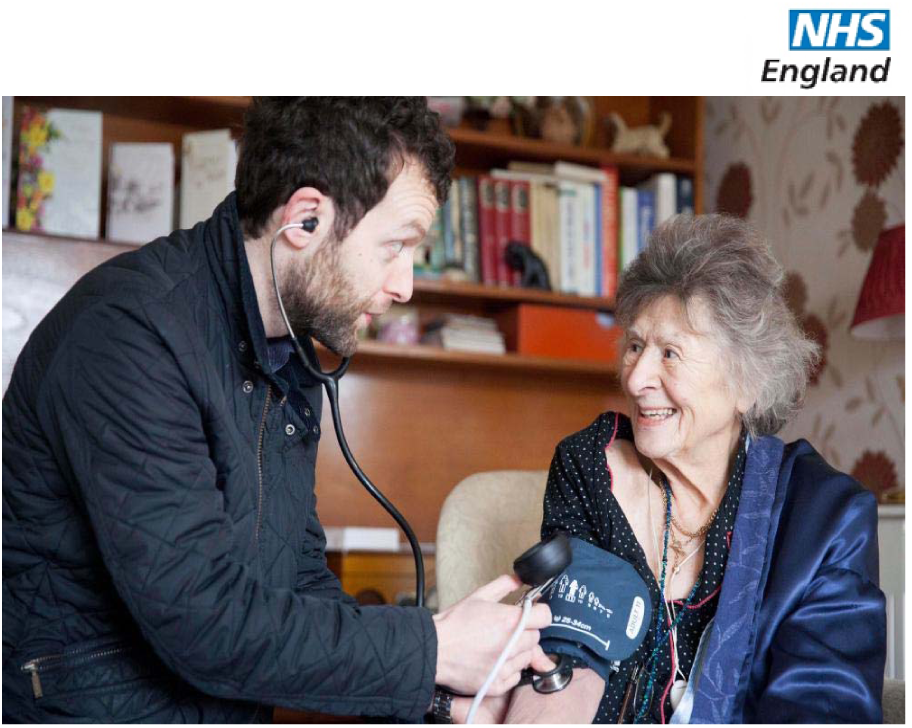

**IF YOU HAVE SYMPTOMS PLEASE COME TO SEE US**

**WE ARE HERE TO LISTEN AND TO HELP**

This leaflet contains some important information about symptoms your doctor would like to see and what you should do if you have them.

**Your guide to making a decision to consult your doctor**

**Why have I received this leaflet?**

Your doctor is encouraging you to visit if you have any concerns or symptoms.

Your doctor would like to reassure you that you will not be wasting our time even if you feel otherwise well.

Your doctor will be supportive and will be happy to discuss your concerns.

**Why are the symptoms mentioned in this leaflet important?**

Occasionally the symptoms mentioned in this leaflet can be a sign of cancer.

Cancer is common, but today 1 in 2 people who are diagnosed with cancer survive.

When cancers are caught earlier the treatment is more successful and has fewer side effects.

Early symptoms can be difficult to spot and may seem minor but if you have any you should check with your doctor.

**Spotting and treating cancer early can save lives**

**Why should I report these symptoms?**

The following symptoms are usually harmless but can be signs of more serious illness, if you have any you should check with

your doctor.

1. **Blood in pee:** You should always report blood in your urine to your doctor.
2. **Blood in poo**: If you see blood when you go to the toilet it is most likely to be piles (haemorrhoids) but you should tell
3. your doctor.
4. **Persistent cough**: You should tell your doctor if your cough gets worse or lasts more than 3 weeks.
5. **Coughing up blood**: If you have coughed up blood, no matter how much or what colour, it is important to tell your
6. doctor.
7. **Difficulty swallowing:** You should check with your doctor if this problem doesn’t go away within a couple of weeks.
8. **Weight loss:** If you lose a noticeable amount of weight without trying to do so, you should tell your doctor.

**Your doctor would like you to tell them about your symptoms**

